# Functional Variation in the *FAAH* Gene is Directly Associated with Subjective Well-being and Indirectly Associated with Problematic Alcohol Use

**DOI:** 10.1101/2023.08.06.23293711

**Authors:** Lisa Bornscheuer, Andreas Lundin, Yvonne Forsell, Catharina Lavebratt, Philippe A. Melas

## Abstract

Fatty acid amide hydrolase (FAAH) is an enzyme that degrades anandamide, an endocannabinoid that modulates mesolimbic dopamine release and, consequently, influences states of well-being. Despite these known interactions, the specific role of FAAH in subjective well-being remains underexplored, particularly with longitudinal data. In our study, we analyzed well-being data collected three years apart using the WHO (Ten) Well-Being Index and genotyped a functional polymorphism in the *FAAH* gene (rs324420, Pro129Thr) in a sample of 2,822 individuals. We found that the A-allele of rs324420, which results in reduced FAAH activity and elevated anandamide levels, was associated with lower well-being scores at both time points. A subsequent phenome-wide association study (PheWAS) validated our well-being findings in the UK Biobank (N=126,132) and revealed an additional association with alcohol dependence. In our cohort, using lagged longitudinal mediation analyses, we uncovered evidence of an indirect association between rs324420 and problematic alcohol use (AUDIT-P) through the pathway of lower well-being. We propose that lifelong elevated anandamide levels can disrupt the endocannabinoid system – a biological contributor to well-being – potentially leading to increased alcohol intake. Further genetic studies and mediation analyses are needed to validate and extend these findings.

## 1. Introduction

Subjective well-being, although complex to define, is a critical construct in understanding human behavior and health. It is typically characterized as a positive evaluation of one’s life and a general sense of contentment (CDC, 2018). While well-being is significantly influenced by societal and social factors such as interpersonal relationships, housing, employment, and institutional trust (Boarini et al., 2012; D’Agostino et al., 2018), there is also evidence supporting a role for specific genes. On average, heritability estimates for various well-being constructs are about 42% (Bartels, 2015). So far, however, single nucleotide polymorphisms (SNPs) account only for roughly 6.3% of this heritability (Baselmans and Bartels, 2018; Jamshidi et al., 2022; Okbay et al., 2016). The complexity in defining and measuring well-being, combined with its moderate heritability estimates, likely contributes to the difficulties in elucidating its underlying genetic architecture. Therefore, insights from preclinical and translational research can provide valuable guidance for studies aiming to identify key genes influencing well-being.

The human *FAAH* gene encodes for the enzyme fatty acid amide hydrolase (FAAH), which degrades N-arachidonoyl ethanolamine (AEA), commonly known as anandamide, a high-affinity partial agonist for the cannabinoid receptor 1 (McKinney and Cravatt, 2005; van Egmond et al., 2021). Anandamide is known to modulate mesolimbic dopamine release (Oleson and Cheer, 2012), a critical component of the reward system involved in the perception of pleasurable experiences (Ferreri et al., 2019) and subjective well-being (Mizrahi et al., 2009; Rutledge et al., 2015). However, the neurobiological effects of anandamide are complex and do not fully mirror those produced by exogenous substances like cannabis (Scherma et al., 2019). For instance, pharmacogenetic studies using either FAAH inhibitors or *Faah* knock-out mice have demonstrated that increased anandamide levels can modulate emotional states without inducing the typical symptoms of cannabinoid intoxication, such as reduced body temperature, catalepsy, or heightened feeding behavior (van Egmond et al., 2021). Moreover, stress has been found to reduce anandamide levels (Yasmin et al., 2020), and pharmacogenetic studies suggest that FAAH inhibition could help regulate stress-related and affective behaviors (Danandeh et al., 2018; Dincheva et al., 2015; Fidelman et al., 2018; Fotio et al., 2023; Mayo et al., 2020).

Despite the recognized relationship between anandamide and well-being, no research to date has examined the link between *FAAH* and subjective well-being within a longitudinally assessed human population. In response to this gap, we utilized data from a population-based cohort assessed at two distinct time points, three years apart, and genotyped a functional genetic variant in the *FAAH* gene, rs324420. This SNP, often referred to as the Pro129Thr variant, results in an amino acid substitution, with the A allele leading to a FAAH variant with reduced expression and activity, which increases anandamide levels (Chiang et al., 2004; Flanagan et al., 2006; Sipe et al., 2002). In addition to our primary research objective, our investigation also led us to examine the potential relationship between rs324420 and problematic alcohol use. Our findings reveal a significant association between the A allele of *FAAH*’s rs324420 and lower well-being, and an indirect association of the same allele with problematic alcohol use via lower well-being. We postulate that lifelong elevated anandamide levels might disrupt the endocannabinoid system, leading to the observed associations later in life.

## 2. Methods

### 2.1. Participants

PART (an acronym from the Swedish ’Psykisk hälsa, Arbete och RelaTioner’), is a longitudinal cohort project designed to investigate the risk and protective factors for mental health in Stockholm County, Sweden (Hällström et al., 2003). The current study utilized rs324420 genotyping data and information extracted from self-administered questionnaires in PART Wave I (1998-2000; 53% response rate, N=10,443) and PART Wave II (2001-2003; 84% response rate, N=8,613) (Bergman et al., 2010; Lundberg et al., 2005). These questionnaires encompassed areas such as demographics, subjective well-being, mental health problems, social support, stressful life events, childhood adversities, and alcohol use, detailed below. The PART project was conducted in accordance with the Code of Ethics of the World Medical Association’s (WMA) Declaration of Helsinki and approved by the ethical review board at Karolinska Institutet. All participants gave informed consent.

### 2.2. Subjective well-being

Subjective well-being, often defined as a person’s cognitive and affective evaluations of their life (Diener et al., 2002), was assessed using the WHO (Ten) Well-Being Index (Bech et al., 1996), a derivative of the WHO (Bradley) Subjective Well-Being Inventory Index (Bradley, 1994). This scale comprises ten items, with a recall period of the previous week. Six items pertain to coping skills and life adjustment (i.e., cognitive evaluations), while the remaining four items focus on symptoms of depression, anxiety, and vitality (i.e., affective evaluations). These items were scored on a scale from ’never’ (0) to ’always’ (3). Higher scores indicate greater well-being, with total scores ranging from 0 to 30. Subjective well-being scores were calculated separately for PART Waves I and II.

### 2.3. Depression and anxiety diagnoses

Depression diagnoses, including major depression, mixed anxiety depression, or dysthymia as per DSM-IV, were identified using the Major Depression Inventory (MDI) supplemented with questions regarding disability due to psychological symptoms (Forsell, 2005; Olsen et al., 2003). These assessment items corresponded to the two weeks preceding questionnaire completion. Anxiety diagnoses, consistent with DSM-IV, were determined using the Sheehan Patient Rated Anxiety Scale (Sheehan, 1983) and the phobia/avoidance component of an instrument by Marks & Mathews (Marks and Mathews, 1979). Even though the American Psychological Association (APA) now classifies obsessive-compulsive disorders (OCD) as a distinct mental health condition, we incorporated it into the anxiety group using screening questions recommended by the Swedish Psychiatric Association and the Swedish Institute for Health Services Development (Wallerblad et al., 2012). This was done to maintain consistency with previous PART publications relying on DSM-IV criteria for an anxiety diagnosis. A history of depression or anxiety was attributed to individuals if symptoms were reported in either PART Wave I or II.

### 2.4. Stressful life events and childhood adversities

Data from PART Waves I and II concerning stressful life events and childhood adversities were extracted and analyzed, as previously described (Bornscheuer et al., 2022; Liu et al., 2015; Melas et al., 2018a; Rahman et al., 2017; Rayman et al., 2020). In brief, stressful life events referred to incidents occurring within 12 months before questionnaire completion, scored based on 28 stressful items including, but not limited to, interpersonal conflicts, separation, severe illness or death of a loved one, significant work-related issues, abortion, and family member victimization. Childhood adversities, occurring before the age of 18, encompassed loss of a parent, parental divorce, serious financial hardships, and severe family conflicts. Both stressful life events and childhood adversities were treated as categorical variables (none or at least 1). Childhood adversity data was obtained from the PART Wave I questionnaire, while stressful life event data were retrieved and separately analyzed for PART Waves I and II.

### 2.5. Social support

The PART questionnaires include two items specifically assessing social support: (i) "There is one person in particular that I feel I can really get support from", and (ii) "Apart from those at home, there are others I can turn to when I’m having a hard time, someone I can easily meet, that I trust and can get real help from when I’m in trouble". Both items were scored on a four-point scale, ranging from "true" to "not true at all". As these items showed strong correlation (Spearman’s rho: 0.477, p<0.001 for PART Wave I and Spearman’s rho: 0.492, p<0.001 for PART Wave II), we consolidated them into a single social support variable, categorized as high (i.e., scoring at or above the mean of both questions) or low (i.e., scoring below the mean). Social support was calculated and analyzed separately for PART Waves I and II.

### 2.6. Alcohol consumption and problematic alcohol use

The Alcohol Use Disorders Identification Test (AUDIT) was utilized to examine alcohol consumption and alcohol-related problems. This tool, previously validated in the PART study (Lundin et al., 2015), comprises 10 items with a total score ranging from 0-40, reflecting drinking habits and associated problems over the past 12 months. AUDIT data from PART Waves I and II were extracted and analyzed in terms of the complete AUDIT scale (items 1-10) and the AUDIT-P scale (items 4-10) that assesses problematic alcohol use.

### 2.7. DNA collection and genotyping

DNA samples were collected from a subset of participants (N=3,018) who responded to PART Waves I and II, using self-administered saliva collection kits (Oragene DNA sample collection kit; DNA Genotek Inc., Canada) as described previously (Melas et al., 2010; Sjoholm et al., 2009). For this study, genotyping of rs324420 was conducted in N=2,915 individuals using a TaqMan SNP genotyping assay on an ABI 7900 HT instrument (Thermo Fisher Scientific, Waltham, MA, USA). N=93 genotyping reactions (3.19%) were ambiguous and were thus excluded from subsequent analyses. We assessed genotyping quality by testing for deviation from the Hardy-Weinberg equilibrium, setting p at < 0.05.

### 2.8. Phenome-wide association study

We employed the Atlas of GWAS Summary Statistics (GWAS Atlas, https://atlas.ctglab.nl/) (Watanabe et al., 2019) to conduct a phenome-wide association study (PheWAS) for rs324420, aiming to validate our findings in distinct cohorts. We set the maximum p-value threshold at 0.01 for this analysis.

### 2.9. Statistical analyses

Normality of the data was assessed using the Shapiro-Wilk test and homoscedasticity was checked with Levene’s test. Spearman’s rank correlation was used to measure correlation between ordinal variables. Differences in subjective well-being scores across groups defined by age, sex, education, social support, mental health diagnosis, stressful life events, and childhood adversities were assessed using the Kruskal-Wallis test. We constructed linear regression models to probe the relationships between rs324420 genotypes, subjective well-being, and AUDIT scores from AUDIT-10 and AUDIT-P. The models explored the crude and adjusted (for age and sex) associations. Multicollinearity was examined using the variance inflation factor (VIF) and tolerance values. Lagged longitudinal mediation analyses were conducted using Model 4 of the PROCESS macro for SPSS (Hayes, 2013) to explore potential indirect associations between rs324420 genotype and AUDIT scores via subjective well-being. We controlled for alcohol use from PART Wave I to remove its potential impact on well-being in the same wave. We also adjusted for potential confounders of the well-being/AUDIT relationship including age, sex, social support, depression, anxiety, stressful life events, and childhood adversities. The indirect effects of rs324420 on AUDIT scores through well-being were estimated using bias-corrected bootstrap confidence intervals based on 5,000 bootstrap samples. All statistical analyses were performed using IBM SPSS Statistics v. 27 (IBM Corp, Armonk, NY, USA) with a significance level set at p<0.05.

## 3. Results

### 3.1. Participant and genotype characteristics

Table 1 illustrates the sociodemographic and diagnostic attributes of genotyped participants, set against subjective well-being scores from PART Wave I. Younger age, female sex, low social support, and a diagnosis of anxiety or depression were significantly associated with lower well-being scores (p<0.001; Table 1). Similarly, lower well-being was significantly associated with experiences of stressful life events in the prior year or reported childhood adversities (p<0.001; Table 1). The rs324420 genotyping results conformed to Hardy–Weinberg equilibrium (chi-square=0.495, p=0.780), with observed genotype frequencies [N=1713 CC carriers (60.7%), N=963 AC carriers (34.12%) and N=146 AA carriers (5.17%)] in line with the overall allele frequencies reported by dbGaP for the total population (C=0.7946, A=0.2053, sample size=374,708; Alpha Allele Frequency release version: 20201027095038).

**Table 1.**
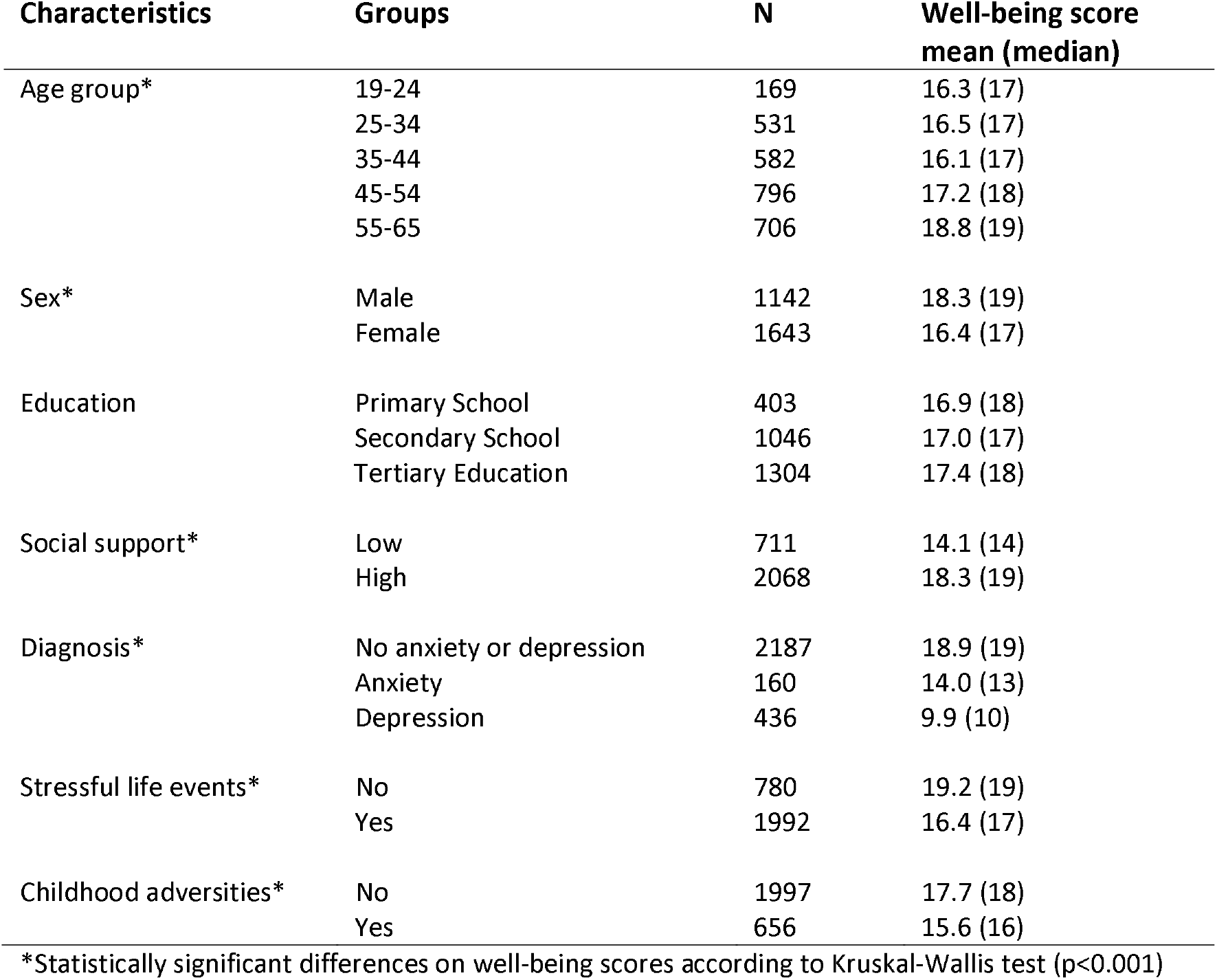
Well-being scores across sociodemographic and diagnostic characteristics of the genotyped study group.

| Characteristics | Groups | N | Well-being score mean (median) |
| --- | --- | --- | --- |
| Age group* | 19-24 | 169 | 16.3 (17) |
|  | 25-34 | 531 | 16.5 (17) |
|  | 35-44 | 582 | 16.1 (17) |
|  | 45-54 | 796 | 17.2 (18) |
|  | 55-65 | 706 | 18.8 (19) |
| Sex* | Male | 1142 | 18.3 (19) |
|  | Female | 1643 | 16.4 (17) |
| Education | Primary School | 403 | 16.9 (18) |
|  | Secondary School | 1046 | 17.0 (17) |
|  | Tertiary Education | 1304 | 17.4 (18) |
| Social support* | Low | 711 | 14.1 (14) |
|  | High | 2068 | 18.3 (19) |
| Diagnosis* | No anxiety or depression | 2187 | 18.9 (19) |
|  | Anxiety | 160 | 14.0 (13) |
|  | Depression | 436 | 9.9 (10) |
| Stressful life events* | No | 780 | 19.2 (19) |
|  | Yes | 1992 | 16.4 (17) |
| Childhood adversities* | No | 1997 | 17.7 (18) |
|  | Yes | 656 | 15.6 (16) |

\*Statistically significant differences on well-being scores according to Kruskal-Wallis test ( $p < 0.001$ )

### 3.2. The FAAH activity-reducing allele is associated with decreased subjective well-being at two distinct time points

Analyzing data from PART Wave I, we identified a significant dose-dependent association between the number of A-alleles at rs324420 (known to reduce FAAH’s expression and activity) and lower subjective well-being. Each additional A-allele was associated with a decrease in well-being scores of 0.54 units (B: -0.54, p<0.01; Table 2). This association remained significant after adjusting for age and sex (B: -0.52, p<0.01; Table 2). A similar significant association was observed in the PART Wave II data, collected three years after Wave I, in both the crude (B: - 0.42, p=0.03; Table 2) and adjusted models (B: -0.41, p=0.03; Table 2). We then evaluated the distinct contributions of the heterozygous (AC) and homozygous risk-allele (AA) genotypes to well-being levels, using the homozygous non-risk allele genotype (CC) as the reference. In the PART Wave I data, individuals homozygous for the A-allele exhibited significantly lower well-being in both the crude (B: -1.29, p=0.01; Table 3) and adjusted models (B: -1.17, p=0.02; Table 3). Heterozygous individuals (AC) also demonstrated a trend towards lower well-being, although this trend did not reach significance in the crude model (B: -0.46, p=0.06) and was at the threshold of significance in the adjusted model (B: -0.47, p=0.05). This pattern, with A-allele homozygosity correlating significantly with lower well-being levels but no significant correlation with heterozygosity, was replicated in the PART Wave II data in both crude (AA genotype, B: - 1.45, p<0.01; Table 3) and adjusted models (AA genotype, B: -1.36, p=0.01; Table 3). Figure 1 graphically displays these findings for PART Wave I (Fig. 1A) and PART Wave II (Fig. 2A), presenting mean predicted well-being scores by genotype group, based on the coefficients from the regression adjusted for age and sex.

**Table 2.**
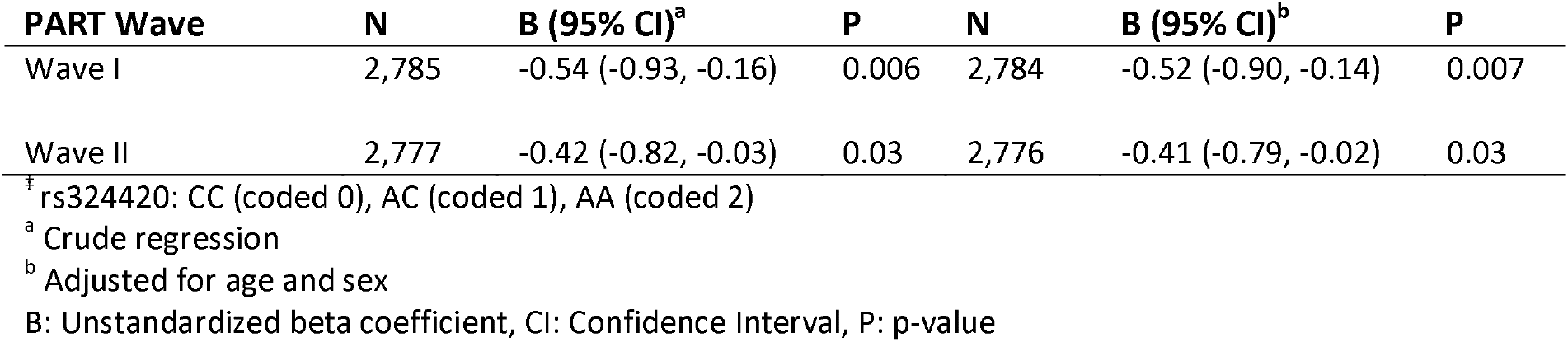
Linear regressions of subjective well-being scores on *FAAH* genotype.

**Table 3.**
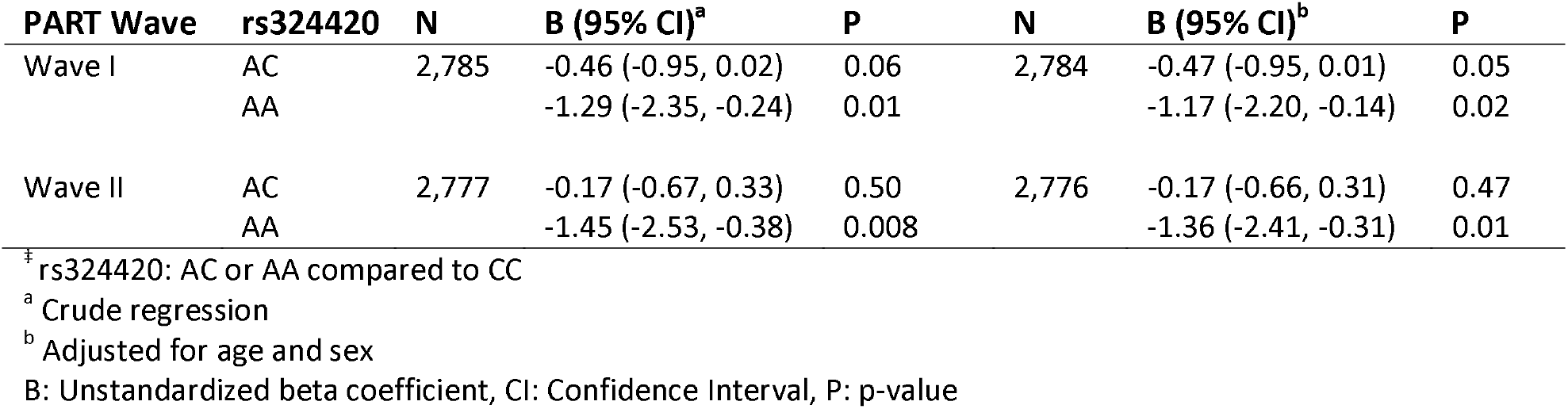
Linear regressions of subjective well-being scores on individual risk-allele *FAAH* genotypes.

**Figure 1.**
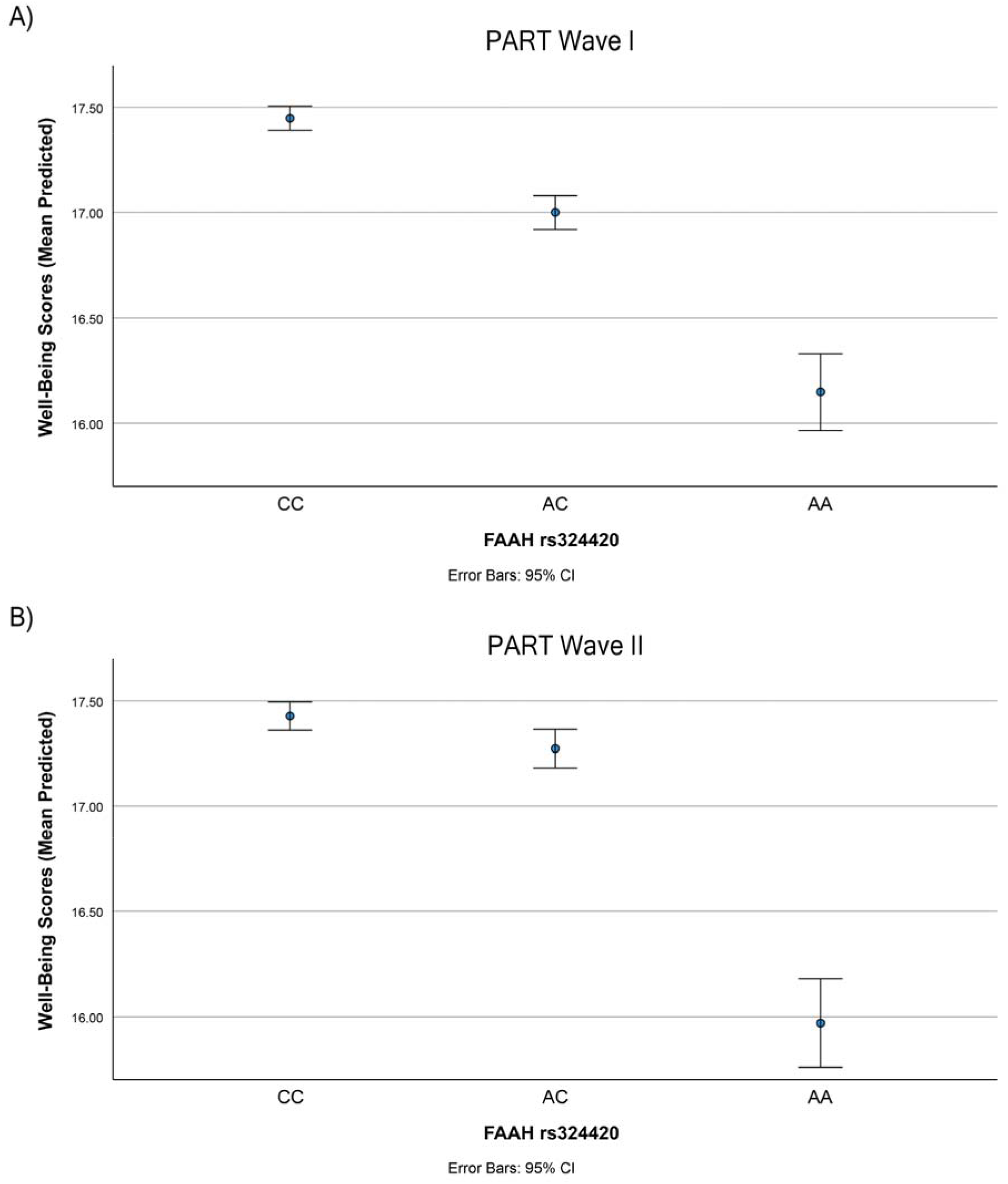
Reduced FAAH activity is associated with lower subjective well-being. The A-allele of rs324420, which encodes threonine, is known to reduce FAAH expression and activity. The linear regression analyses conducted on subjective well-being against rs324420 revealed that well-being scores were significantly lower in A-homozygotes compared to C-homozygotes (Table 3). The graph illustrates the relationship between the three genotype groups of rs324420 and their respective predicted well-being scores in (A) PART Wave I and in (B) PART Wave II, calculated using coefficients from the regressions adjusted for age and sex. The error bars represent the 95% confidence intervals (CI).

### 3.3. Validation of the genetic association between *FAAH*’s rs324420 and subjective well-being

To corroborate our findings across different cohorts, we performed a Phenome-Wide Association Study (PheWAS) for rs324420, encompassing 4,756 GWASs and identifying 66 traits significantly linked to rs324420 (P<0.01; Table S1). Within the psychiatric domain, the PheWAS unveiled three significant traits, the first of which affirmed our well-being results: (a) Happiness and subjective well-being (Effect allele: C, N=126,132, p=0.008, Cohort: UK Biobank), (b) Nap during day (Effect allele: A, N=386,124, p=0.00006, Cohort: UK Biobank), and (c) Alcohol dependence (Effect allele: A, N=1,161, p=0.005, Cohort: the Study of Addiction, Genetics and Environment; SAGE).

### 3.4. Indirect Association between *FAAH*’s rs324420 and Problematic Alcohol Use Through Lower Subjective Well-Being

The PheWAS results within the psychiatric domain suggested an association between the A-allele of rs324420 and alcohol dependence. To further investigate this relationship, we used AUDIT data from our cohort. However, we did not observe a significant association between rs324420 and AUDIT-10 or AUDIT-P (Table S2). This inconsistency led us to consider the possibility of an indirect relationship between the A-allele of rs324420 and increased alcohol use, potentially facilitated through lower subjective well-being. To explore this hypothesis, we conducted a lagged longitudinal mediation analysis using well-being scores from PART Wave I and AUDIT data from PART Wave II. This analysis showed no direct association between the *FAAH* genotype and the two AUDIT scales, but did reveal significant indirect associations via lower well-being (Table 4). However, upon controlling for alcohol use levels from PART Wave I (to remove alcohol’s potential impact on well-being in the same wave) and other potential confounders from PART Wave II, significant indirect associations persisted for AUDIT-P only (Table 4). Taken together, these findings support the hypothesis that decreased FAAH activity may indirectly contribute to problematic alcohol use through its impact on well-being.

**Table 4.** Mediation analyses of the *FAAH*^‡^ genotype’s effect on alcohol use through well-being.

| AUDIT scale | N | Direct effect<br>(Boot 95% CI) | Sig | Indirect effect<br>(Boot 95% CI) | Sig |
| --- | --- | --- | --- | --- | --- |
| AUDIT-10<sup>a</sup> | 2,622 | -0.096 (-0.285, 0.092) | No | 0.033 (0.007, 0.064) | Yes |
| AUDIT-10<sup>b</sup> | 2,550 | -0.082 (-0.212, 0.047) | No | 0.004 (-0.002, 0.015) | No |
| AUDIT-10<sup>c</sup> | 2,411 | -0.044 (-0.179, 0.091) | No | 0.002 (-0.003, 0.011) | No |
| AUDIT-P<sup>a</sup> | 2,628 | 0.006 (-0.107, 0.119) | No | 0.030 (0.007, 0.056) | Yes |
| AUDIT-P<sup>b</sup> | 2,555 | 0.019 (-0.071, 0.110) | No | 0.015 (0.003, 0.030) | Yes |
| AUDIT-P<sup>c</sup> | 2,416 | 0.036 (-0.058, 0.131) | No | 0.005 (0.0003, 0.012) | Yes |

<sup>‡</sup> rs324420: CC (coded 0), AC (coded 1), AA (coded 2)
<sup>a</sup> Crude model
<sup>b</sup> Model adjusted for AUDIT-10 in PART Wave I
<sup>c</sup> Model adjusted for AUDIT-10 in PART Wave I, age, sex, social support in PART Wave II, depression, anxiety, stressful life events in PART Wave II, childhood adversities
AUDIT-10: Full AUDIT scale (10 items), AUDIT-P: Alcohol problem items
Boot CI: Bootstrap Confidence Interval, Sig: Statistical significance

## 4. Discussion

Our study revealed a significant dose-dependent association between the minor (A; Thr) allele of the rs324420 genetic variant, a mutation that downregulates the expression and activity of FAAH—the enzyme primarily responsible for breaking down the endocannabinoid anandamide—and lower subjective well-being, as measured by the standardized WHO (Ten) Well-Being Index. This observation held across two measurement periods, three years apart, with each additional A-allele associated with a decrease in well-being scores. Importantly, these findings were not only observed in crude models but also remained significant after adjusting for age and sex, and further validation was obtained through a PheWAS analysis using GWAS data from a distinct and substantially larger cohort—the UK Biobank. Our investigation also illuminated a potential connection between rs324420 and alcohol consumption. Specifically, lagged longitudinal mediation analyses revealed significant indirect associations between the *FAAH* genotype and AUDIT scores via lower well-being. However, when controlling for alcohol use levels from PART Wave I and other potential confounders, the indirect associations persisted only for problematic alcohol use, as measured by AUDIT-P. These findings highlight the potential for decreased FAAH activity to indirectly contribute to problematic alcohol use through its impact on well-being

Interestingly, the only preceding study to associate the *FAAH* gene with well-being utilized population genetic data from an allele frequency database, correlating the allele frequencies of rs324420 with national averages of self-reported happiness (Minkov and Bond, 2017). This study identified an inverse association to ours, with the A-allele linked to increased happiness. This contrasting finding is likely attributed to the different methodologies employed: the previous study relied on an allele frequency database, while ours employed direct genotyping of the study group. Our results are further substantiated by replication efforts using a PheWAS, which once again linked the A-allele with reduced well-being and happiness in a GWAS employing the UK Biobank (Watanabe et al., 2019). Additionally, evidence points to the A-allele of rs324420 as a potential risk factor for anxiety and depression (Lazary et al., 2016), which also aligns with our genetic findings. Notably, individuals homozygous for the C allele of rs324420 have been found to exhibit more positive affective states post-placebo administration (Pecina et al., 2014), and increased happiness following cannabis use (Schacht et al., 2009).

While our findings demonstrate a robust association, it is noteworthy that no GWAS to date has linked *FAAH* to well-being at a genome-wide significance level (i.e., p<5×10^−8^). Most GWAS studies of well-being have relied on single-item measures or well-being indices not derived from validated instruments, which might have limited their ability to detect this association. Nonetheless, several GWAS studies examining well-being through different measures have succeeded in identifying several genome-wide significant variants (Okbay et al., 2016; Baselmans and Bartels, 2018; Baselmans et al., 2019; Jamshidi et al., 2022). One recent study attempting to leverage all available well-being GWAS data to prioritize and functionally annotate variants of importance, identified three genes—*PSMC3*, *ITIH4*, and *SERPINC1*—as probable candidates in the regulation of the well-being spectrum (Pyne et al., 2021). In light of our findings, which were obtained using the standardized WHO (Ten) Well-Being Index and further supported by a PheWAS analysis, we propose that *FAAH* may also be a significant contributor to well-being. If this is the case, *FAAH* may emerge as a genome-wide significant locus in future, larger-scale GWAS studies on well-being, particularly those employing more comprehensive and validated measures of well-being.

In our study, we also found evidence of an indirect association between *FAAH*’s rs324420 and problematic alcohol use, potentially mediated by a decrease in well-being. Prior research has suggested a link between FAAH and various substance use disorders, including alcohol and other drugs (Best et al., 2022; Best et al., 2021; Best et al., 2020; Buhler et al., 2014; Flanagan et al., 2006; Melroy-Greif et al., 2016; Sipe et al., 2002; Sloan et al., 2018; Zhang et al., 2020), although conflicting findings have been noted regarding alcohol use (Tyndale et al., 2007) and rs324420’s effect allele (Buhler et al., 2014). Our PheWAS analysis corroborated these links, but interestingly, we did not identify a direct association between rs324420 and alcohol use in our cohort. This discrepancy could be due to our study being underpowered. However, the largest GWAS to date on alcohol use phenotypes, including alcohol use disorder (Kranzler et al., 2019), problematic alcohol use (Zhou et al., 2020) and drinks per week (Saunders et al., 2022), have also not found genome-wide associations with *FAAH*. This suggested to us that the relationship between *FAAH*’s rs324420 and substance use may be indirect, via reduced well-being that prompts substance use. Indeed, our lagged longitudinal mediation analysis using well-being scores from PART Wave I and AUDIT data from PART Wave II revealed significant indirect associations through well-being. These associations held, specifically for problematic alcohol use (AUDIT-P), even after controlling for potential confounders, including baseline alcohol use. This aligns with previous studies that have suggested the relationship between the rs324420 A-allele and drinking outcomes might be mediated by coping motives (Best et al., 2021). Specifically, the *FAAH*’s rs324420 AC/AA genotype group showed a significant indirect effect indicating a higher propensity to drink to alleviate negative mood or to escape worries (Best et al., 2021). Taken together, these data reinforce the idea that lower FAAH activity might instigate excessive alcohol consumption by impacting psychological aspects of negative reinforcement, a key factor implicated in substance use disorders (Koob, 2022).

Although our study provides robust associations, it is primarily limited by its correlational nature, preventing us from providing direct mechanistic insights into how FAAH’s reduced activity contributes to the observed associations. Preclinical research with *Faah* knockout mice has lent credence to the idea that behavioral changes brought on by increased anandamide levels in the brain mainly transpire through the CB1 receptor (Cravatt et al., 2001). Additionally, studies using CB1 knockouts have suggested a receptor/ligand interplay between CB1 and anandamide, where tonic activation of the receptor seems to govern the biosynthesis of this endogenous ligand (Di Marzo et al., 2000). Insights into the effects of FAAH and anandamide could also be drawn from cannabis studies, given that its psychoactive component, tetrahydrocannabinol (THC), also activates the CB1 receptor. Previous human research has linked frequent teenage cannabis use to poorer functional well-being and problematic substance use (Shanahan et al., 2021). Similarly, animal studies have consistently identified prenatal and adolescent neurodevelopmental periods as crucial to understanding the detrimental behavioral and molecular effects of exogenously administered cannabinoids (Kononoff et al., 2018; Melas et al., 2018b; Qvist et al., 2022; Scherma et al., 2020; Tortoriello et al., 2014). Consequently, we posit that FAAH may play a key role in modulating well-being through its influence on anandamide levels, which act on cannabinoid receptors crucial for healthy neurodevelopment (Basavarajappa et al., 2009). However, it’s important to note that FAAH’s enzymatic activity extends beyond anandamide to include a broader array of fatty acid amides, including other N-acylethanolamines (van Egmond et al., 2021), oleamide (Cravatt et al., 1996), and N-acyltaurines (Sasso et al., 2016). Anandamide also goes beyond interacting solely with cannabinoid receptors; it serves as a full agonist of the transient receptor potential vanilloid type-1 (TRPV1) ion channel (Zygmunt et al., 1999) and can activate the nuclear receptor peroxisome proliferator-activated receptor γ (PPARγ) (Bouaboula et al., 2005). Given these additional downstream pathways, future investigations into FAAH’s influence on well-being should consider all these potential interactions.

In conclusion, our study provides compelling genetic evidence supporting FAAH’s role in subjective well-being. This association was found to be significant at two distinct time points within the same cohort and was further corroborated in an independent cohort. To the best of our knowledge, we also present the first evidence of a possible indirect association between *FAAH* and alcohol use through well-being. Based on our results, we propose that genetically driven elevation of anandamide levels, inherent in carriers of FAAH’s rs324420 A-allele, might disrupt the endocannabinoid system from conception onwards. This disruption could increase the likelihood of diminished well-being, which in turn could lead to problematic alcohol consumption. However, more genetic studies and mediation analyses are necessary to validate our findings. Furthermore, additional translational and mechanistic investigations are required to comprehend the role of cannabinoid compounds and their target receptors in shaping complex psychological constructs such as well-being.

## Data Availability

All data produced in the present study are available upon reasonable request to the authors.

## Author contribution statement

Lisa Bornscheuer: Investigation; Methodology; Writing - review & editing. Andreas Lundin: Methodology; Supervision; Writing - review & editing. Yvonne Forsell: Methodology; Project administration; Resources; Writing - review & editing. Catharina Lavebratt: Funding acquisition; Methodology, Resources; Supervision; Writing - review & editing. Philippe A. Melas: Conceptualization; Data curation; Formal analysis; Funding acquisition; Investigation; Methodology; Project administration; Resources; Supervision; Validation; Visualization; Writing - original draft.

## Declaration of Generative AI and AI-assisted technologies in the writing process

Statement: During the preparation of this work the authors used ChatGPT (Open AI, San Francisco, CA, ISA) in order to improve readability and language, and to get feedback on the use of statistical software. After using this tool, the authors reviewed and edited the content as needed and take full responsibility for the content of the publication.

