## Supplemental Table S1 for "Functional Variation in the *FAAH* Gene is Directly Associated with Subjective Well-being and Indirectly Associated with Problematic Alcohol Use"

**Supplemental Table S1. Phenome-Wide Association Study (PheWAS) identified 66 traits significantly linked to *FAAH*’s rs324420 at p<0.01. Within the psychiatric domain, the PheWAS unveiled three significant traits (highlighted in yellow).**

| **Domain** | **Trait** | **P-value** | **N** | **EA** |
| --- | --- | --- | --- | --- |
| Activities | Number of days/week of vigorous physical activity 10+ minutes | 0.003794 | 368164 | C |
| Activities | Time spent driving | 0.005381 | 258336 | A |
| Activities | Types of physical activity in last 4 weeks: Other exercises (eg: swimming, cycling, keep fit, bowling) | 0.001304 | 384450 | C |
| Activities | Positive affect (univariate) | 0.009076533 | 410603 | A |
| Activities | Strenuous sports or other exercises | 0.0047 | 350492 | C |
| Activities | Vigorous physical activity | 0.0066 | 261055 | C |
| Activities | Moderate to vigorous physical activity levels | 0.00074 | 377234 | C |
| Cardiovascular | Heart rate | 0.002564 | 85787 | A |
| Cardiovascular | Pulse rate (automated reading) | 0.0006695 | 361411 | A |
| Cardiovascular | Heart rate recovery at 40 secnds | 0.0069 | 58818 | C |
| Cardiovascular | Posterior wall thickness | 0.009263 | 19373 | A |
| Cardiovascular | Left ventricular mass | 0.003373 | 19076 | A |
| Cardiovascular | Resting heart rate | 0.0000085 | 458969 | A |
| Dermatological | Male-specific factors - Hair/balding pattern: Pattern 1 | 0.001682 | 176380 | C |
| Dermatological | Male pattern baldness (BOLT LMM infinitesimal mixed model) | 0.0072 | 205327 | A |
| Dermatological | Male pattern baldness (BOLT LMM non-infinitesimal mixed model) | 0.0087 | 205327 | A |
| Environment | Illnesses of mother: Breast cancer | 0.006447 | 367939 | C |
| Gastrointestinal | Ulcerative colitis | 0.005761 | 45975 | A |
| Gastrointestinal | Non-cancer illness code, self-reported: gastro-oesophageal reflux (gord) / gastric reflux | 0.002621 | 289307 | A |
| Immunological | CD123 on 11c+123+DC | 0.004897054 | 669 | A |
| Metabolic | Amino acid::Glycine, serine and threonine metabolism::threonine | 0.009639 | 6020 | A |
| Metabolic | Amino acid::Histidine metabolism::histidine | 0.00237 | 7804 | A |
| Metabolic | Lipid::Bile acid metabolism::glycochenodeoxycholate | 0.003626 | 7087 | A |
| Metabolic | Lipid::Carnitine metabolism::carnitine | 0.009458 | 7797 | A |
| Metabolic | Lipid::Essential fatty acid::docosahexaenoate (DHA; 22:6n3) | 0.001206 | 7818 | C |
| Metabolic | Lipid::Essential fatty acid::linolenate [alpha or gamma; (18:3n3 or 6)] | 0.006324 | 7786 | C |
| Metabolic | Lipid::Fatty acid metabolism (also BCAA metabolism)::propionylcarnitine | 0.002213 | 7813 | A |
| Metabolic | Lipid::Long chain fatty acid::stearidonate (18:4n3) | 0.001075 | 7775 | C |
| Metabolic | Lipid::Lysolipid::1-arachidonoylglycerophosphoethanolamine* | 0.00762 | 7798 | A |
| Metabolic | Lipid::Lysolipid::1-heptadecanoylglycerophosphocholine | 0.006804 | 7422 | C |
| Metabolic | Peptide::gamma-glutamyl::gamma-glutamylvaline | 0.0009626 | 7753 | A |
| Metabolic | ::::X-06126 | 0.005869 | 7785 | A |
| Metabolic | ::::X-11261 | 0.004773 | 7771 | C |
| Metabolic | ::::X-11374 | 0.001753 | 2609 | A |
| Metabolic | ::::X-11478 | 0.008187 | 6593 | C |
| Metabolic | ::::X-11529 | 0.005358 | 6664 | A |
| Metabolic | ::::X-11792 | 0.002144 | 2442 | C |
| Metabolic | Peptide::Dipeptide::X-12244--N-acetylcarnosine | 0.001479 | 6608 | C |
| Metabolic | ::::X-12556 | 0.006334 | 7483 | A |
| Metabolic | ::::X-12729 | 0.006026 | 1753 | C |
| Metabolic | 22:6, docosahexaenoic acid (DHA) | 0.008215 | 13499 | C |
| Metabolic | OmegaL3 fatty acids | 0.001872 | 13544 | C |
| Metabolic | Adiponectin | 0.004142 | 7825 | C |
| Metabolic | 25-Hydroxyvitamin D level | 0.008131 | 79366 | C |
| Neurological | Inferior fronto-occipital fasciculus fractional anisotropy | 0.001681 | 17706 | A |
| Neurological | Splenium of corpus callosum fractional anisotropy | 0.008783 | 17706 | A |
| Neurological | External capsule mean diusivities | 0.009321 | 17706 | C |
| Neurological | Inferior fronto-occipital fasciculus mean diusivities | 0.001051 | 17706 | C |
| Neurological | External capsule radial diusivities | 0.009843 | 17706 | C |
| Neurological | Inferior fronto-occipital fasciculus radial diusivities | 0.0001234 | 17706 | C |
| Nutritional | Lamb/mutton intake | 0.009147 | 384188 | A |
| Nutritional | Bread intake | 0.002949 | 377627 | A |
| Nutritional | Never eat eggs, dairy, wheat, sugar: Sugar or foods/drinks containing sugar | 0.002323 | 384986 | C |
| Nutritional | Never eat eggs, dairy, wheat, sugar: I eat all of the above | 0.001757 | 384986 | A |
| Psychiatric | Alcohol dependence | 0.0050206 | 1161 | A |
| Psychiatric | Nap during day | 0.00006174 | 386124 | A |
| Psychiatric | Happiness and subjective well-being - General happiness | 0.008221 | 126132 | C |
| Reproduction | Had menopause (female) | 0.009168 | 175519 | A |
| Reproduction | Dysmenorrhea pain severity | 0.006661616 | 11348 | C |
| Reproduction | Menstruation quality of life impact: Increased appetite | 0.004765989 | 11348 | C |
| Skeletal | Total-body less head BMD | 0.00412804 | 10414 | A |
| Skeletal | Total-body lean mass | 0.0092822 | 10414 | A |
| Skeletal | Total-body less head BMD and total body lean mass (bivariate meta-analysis) | 0.00515213 | 10414 | NA |
| Skeletal | Osteoarthritis of hip or knee (hospital diagnosed) | 0.00577852 | 32970 | C |
| Skeletal | Heel bone mineral density | 0.0056 | 394929 | C |
| Social Interactions | Number of full brothers | 0.00537 | 380062 | A |
