## Supplemental Table S2 for "Functional Variation in the *FAAH* Gene is Directly Associated with Subjective Well-being and Indirectly Associated with Problematic Alcohol Use"

**Supplemental Table S2. Linear regression of alcohol use on FAAH genotype^‡^**

| **PART Wave** | **AUDIT scale** | **N** | **B (95% CI)^a^** | **P** | **B (95% CI)^b^** | **P** |
| --- | --- | --- | --- | --- | --- | --- |
| Wave I | AUDIT-10 | 2,647 | 0.02 (-0.16, 0.21) | 0.78 | 0.07 (-0.11, 0.25) | 0.45 |
|  | AUDIT-P | 2,653 | 0.04 (-0.06, 0.16) | 0.42 | 0.05 (-0.05, 0.17) | 0.30 |
| Wave II | AUDIT-10 | 2,644 | -0.07 (-0.26, 0.11) | 0.45 | -0.03 (-0.21, 0.15) | 0.72 |
|  | AUDIT-P | 2,651 | 0.02 (-0.08, 0.14) | 0.64 | 0.04 (-0.07, 0.15) | 0.48 |

^‡^ rs324420: CC (coded 0), AC (coded 1), AA (coded 2)

^a^ Crude regression

^b^ Adjusted for age and sex
